# Consumer Perspectives on the Australian Rheumatology Association Rheumatoid Arthritis Clinical Care Standard

**DOI:** 10.1101/2025.08.11.25333417

**Authors:** Gabriella R. Venter, Rosemary Ainley, Angela Brown, Shirani Wright, Maria B. Sukkar, Catherine L. Hill

## Abstract

**Objective:** The Australian Rheumatology Association released Australia’s first Rheumatoid Arthritis (RA) Clinical Care Standard in November 2023, outlining twelve quality statements to improve RA care. Our study explores consumer perspectives on these quality statements to understand care priorities and inform the development of corresponding quality care indicators.

**Methods:** In the development of the Standard, a national online survey was promoted via Australian professional and consumer arthritis organisations to establish consensus with quality statements. Free-text comments from consumers were thematically analysed using NVivo 14. Related statements were grouped to identify overlapping themes, supportive care statements were analysed individually, and consumer agreement was obtained.

**Results:** 605 consumer responses were received (585 people living with RA, 20 carers). Over half (N=323, 53.4%) provided free-text comments, with 1377 comments received. Key themes in pain management included poor control, the complexities of analgesia, and impacts on quality of life. For emotional and psychological wellbeing, barriers to support and the connection between physical and psychosocial wellbeing were highlighted. Self-management, barriers encountered, and benefits experienced were discussed for both physical activity and supportive resources. Delays in diagnosis or treatment and difficulty accessing rheumatologists were described for accessing specialist care. Medication concerns, shared decision-making, and disease control were key themes in disease and medication management. Consumers cited poor understanding of risks, inadequate care, and confusion about responsibility for preventative healthcare.

**Conclusion:** Our findings revealed several barriers, enablers, and impacts experienced by consumers and demonstrate that consumers value holistic and team-based care, effective communication, shared decision-making, and self-management.

## INTRODUCTION

Rheumatoid arthritis (RA) is an autoimmune inflammatory condition primarily affecting the joints which can result in significant pain, deformity, and impaired function. In 2022, RA was estimated to affect 514,000 people in Australia (2% of the population) and contributed to 5.1 deaths per 100,000 population^1^. The management of RA involves disease modifying anti- rheumatic drugs (DMARDs) to suppress the immune system and control inflammation. Early diagnosis and treatment of RA improves outcomes, ultimately reducing disability and deformity^2,3^.

To improve the quality of care for people living with RA in Australia, the Australian Rheumatology Association (ARA) developed the first national RA Clinical Care Standard for diagnosis and management of RA, released in November 2023^4^. The Standard was created by an interprofessional working group with consumer representation using a series of facilitated workshops and was informed by clinical practice guidelines, international quality criteria, and consumer recommendations^5^. It consists of twelve quality statements to improve the quality of care for people living with RA and is centred around seven guiding principles: high-quality care, culturally responsive care, holistic care, team-based care, effective communication, shared decision-making, and self-management^4^. The Standard serves as an important benchmark for healthcare providers and provides consumers with guidance as to the care that they can expect.

In addition to more traditional biomedical healthcare models, the benefits of a holistic and patient-centred approach are increasingly recognised^6–8^. Acknowledging that most RA management is driven by the patient in their own environment and without input from healthcare professionals, there has been a recent focus on the importance of involving patients and carers as key members of the healthcare team^8^. This approach has been shown to improve health outcomes, acceptability of and adherence to recommended management plans, and satisfaction with care^6^.

As part of the development of the RA Clinical Care Standard, we used a national online survey to explore consumer opinions and experiences regarding the quality statements. By focusing on consumer viewpoints, we hope to develop an understanding of consumer priorities and identify key areas to address to improve the quality and experiences of care for people living with RA in Australia. Our findings will also be used to inform the development of RA quality care indicators to measure the care described in the Standard.

## METHODS

### STUDY DESIGN

As part of the development of the ARA RA Clinical Care Standard, a national online survey was conducted to seek consumer and healthcare provider consensus or non-consensus with the proposed quality statements. The survey included collection of free-text comments which were analysed to identify common themes.

### DATA COLLECTION

Consumers and healthcare professionals were invited to participate in a national online survey using Research Electronic Data Capture (REDCap) software. The survey was promoted across multiple consumer and professional arthritis organisations and was disseminated via multiple communication strategies; this included direct e-mail to members, newsletter articles, social media and articles in specialist publications.

Consumers were asked to indicate whether they were a carer or a person living with RA, and relevant demographic information was collected. All responses were anonymous.

The survey presented consumers with seven guiding principles and 13 proposed quality statements aiming to drive priority quality improvement in RA care. The rationale for each statement and links to supporting references were also provided. Consumers were asked to indicate how important each statement was for improving the quality of RA care (priority), as well as their agreement with the content of the statement; these findings are presented elsewhere^5^. Consumers were additionally invited to provide free-text comments on the content and wording of each statement.

### DATA ANALYSIS

Free-text comments were thematically analysed and NVivo14 was used to assist with coding of themes. Comments with only one to two words (e.g. agree, no, yes, no comment) were excluded from the analysis. Related quality statements were grouped according to areas of care (Figure 1), and a multi-step approach to thematic analysis was applied^9,10^. Analysis was performed by two researchers (GV and CH) until consensus about key themes was achieved.

**Figure 1:**
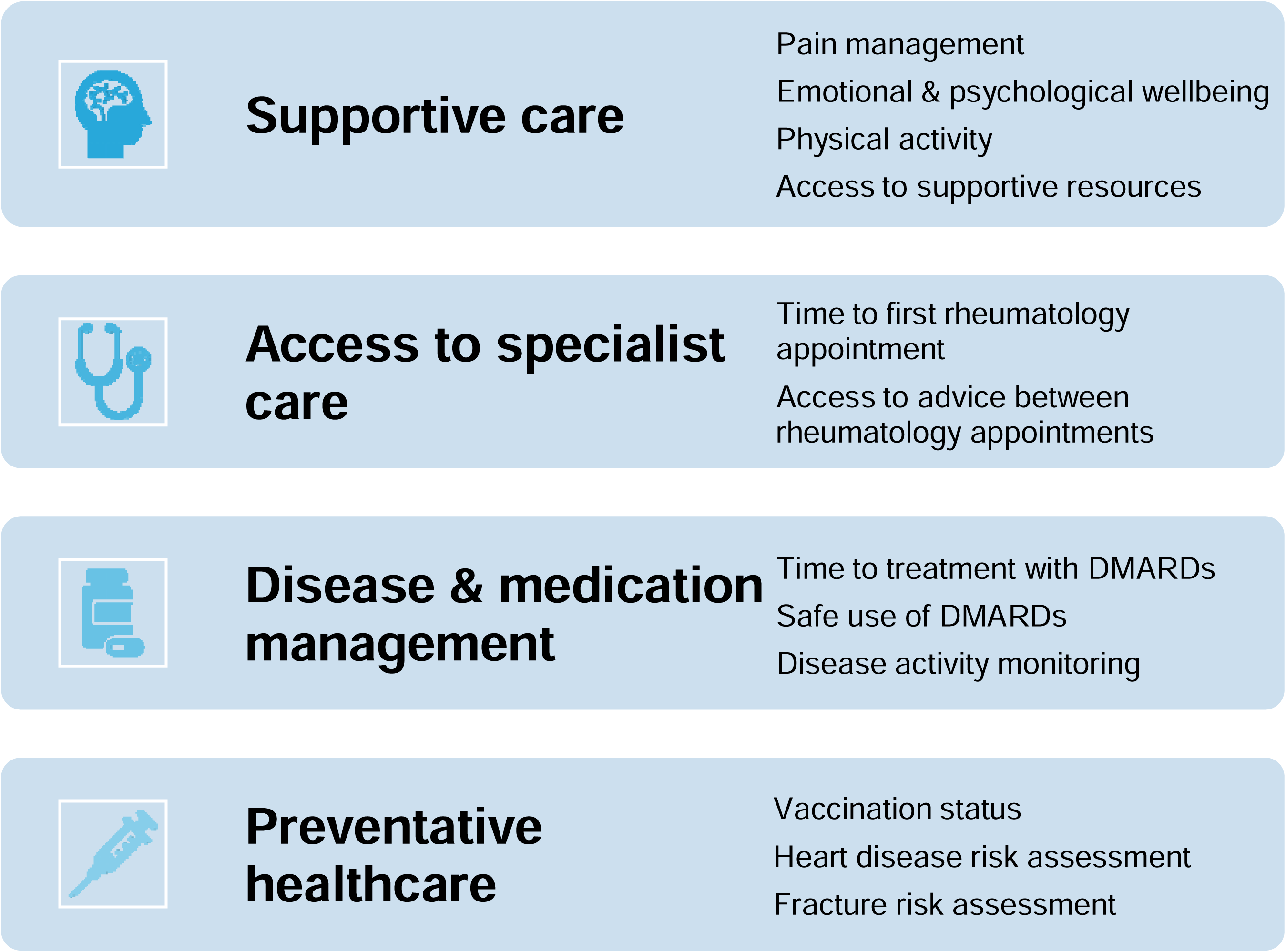
Grouping of Related Quality Statements.

An initial inductive analysis was performed for data familiarisation and to identify common themes for each quality statement. This was subsequently used to inform a framework analysis to identify overlapping themes for grouped statements in each area of care.

After statements were grouped and initial analysis was performed, the findings were presented to three consumers (RA, AB, SW) to seek their input on the approach used. Consumers highlighted the importance of supportive care statements in understanding the patient experience, particularly pain management, and suggested that these should be analysed individually to preserve the richness of the findings.

### CONSUMER PARTICIPATION IN RESEARCH

Consumers with lived experience of RA were involved in all aspects of this study, from study conception and design to thematic analysis, and are authors of this manuscript (RA, AB, SW). The quality of consumer engagement was very high, with all three consumers indicating they strongly agreed with the 22 statements in the Patient Engagement in Research Scale (PIERS-22)^11^.

### ETHICS APPROVAL

Ethical approval for this study was obtained from Central Adelaide Local Health Network Human Research Ethics Committee (No. 17784).

## RESULTS

The survey was open for 6 weeks from 9 August until 20 September 2023. A total of 605 consumer responses were received: 585 from people living with RA and 20 from carers.

Respondents living with RA were mostly female (n=471, 80.5%) and over 55 years of age (n=419, 71.6%). About one third (n=180, 30.8%) were diagnosed within the last 5 years, and 61.5% (n=360) were over 45 years of age at the time of diagnosis. The geographic spread of consumers was representative of the Australian population, with 54.4% (n=318) living in metropolitan areas and 12.6% (n=74) in rural or remote areas. Sixteen respondents (2.7%) identified as Aboriginal and/or Torres Strait Islander people, and 6.2% (n=36) spoke a language other than English at home.

Over half (n=323, 53.4%; 310 people living with RA and 13 carers) provided at least one free-text comment, and demographic information was similar for those who provided free-text comments and those who did not (Table 1). After exclusion of one- or two-word comments, a total of 1377 responses were received (Table 2).

**Table 1:**
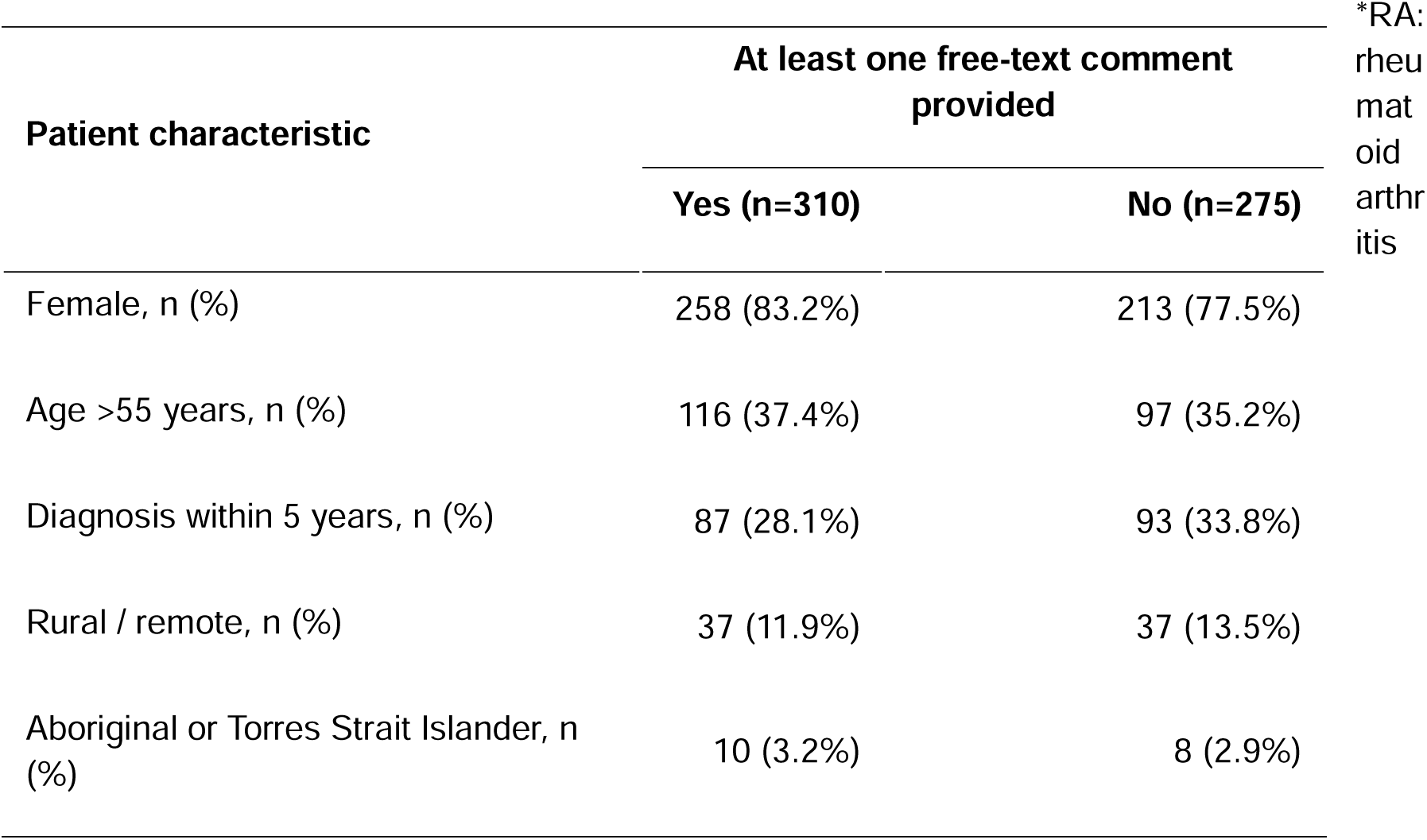
Demographic information for people living with RA* who provided free-text comments vs those who did not.

**Table 2:** Number of free-text comments received for each quality statement.

| Quality Statement by Group |  | Comments received (N) |
| --- | --- | --- |
| Supportive care |  | 500 |
|  | Pain management | 133 |
|  | Emotional and psychological wellbeing | 132 |
|  | Support to engage in physical activity | 160 |
|  | Access to supportive resources | 75 |
| Access to specialist care |  | 287 |
|  | Time to first rheumatology appointment | 134 |
|  | Access to advice between rheumatology appointments | 153 |
| Disease & medication management |  | 300 |
|  | Time to treatment with DMARDs* | 126 |
|  | Safe use of DMARDs* | 68 |
|  | Disease activity monitoring | 106 |
| Preventative healthcare |  | 290 |
|  | Vaccination status | 102 |
|  | Heart disease risk assessment | 107 |
|  | Fracture risk assessment | 81 |
| Total |  | 1377 |

\*DMARDs: disease modifying anti-rheumatic drugs

### THEMATIC ANALYSIS OF QUALITY STATEMENTS RELATED TO SUPPORTIVE CARE

#### Illustrative quotes are presented in Table 3

##### PAIN MANAGEMENT

Inadequate pain control was frequently referenced, and many consumers reported that their pain management was not addressed or insufficient. This resulted in feeling unheard and unsupported, and had significant negative impacts on mental health, social participation, and emotional wellbeing. Pain was also felt to impair function, independence, and the ability to work.

**Table 3:**
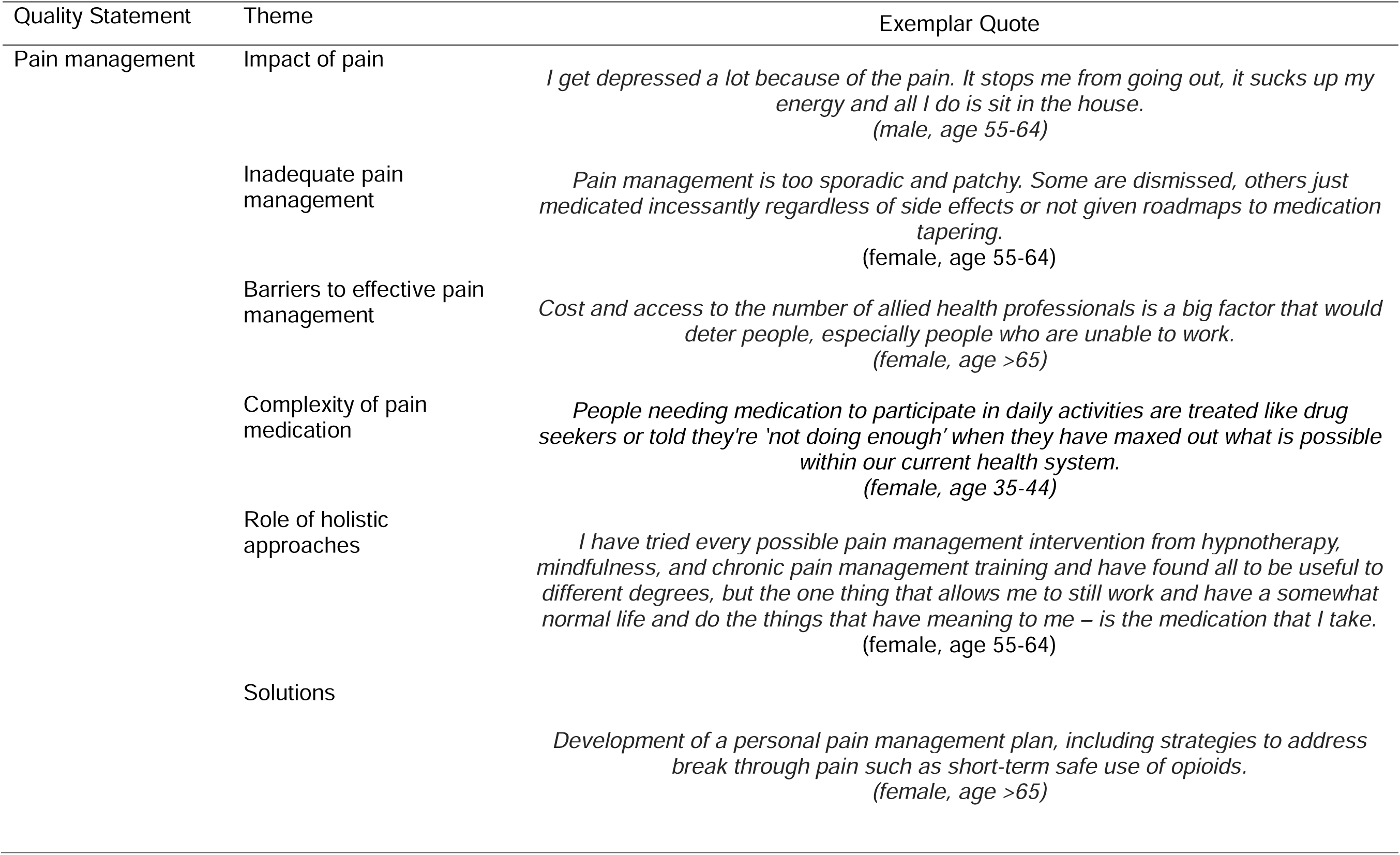
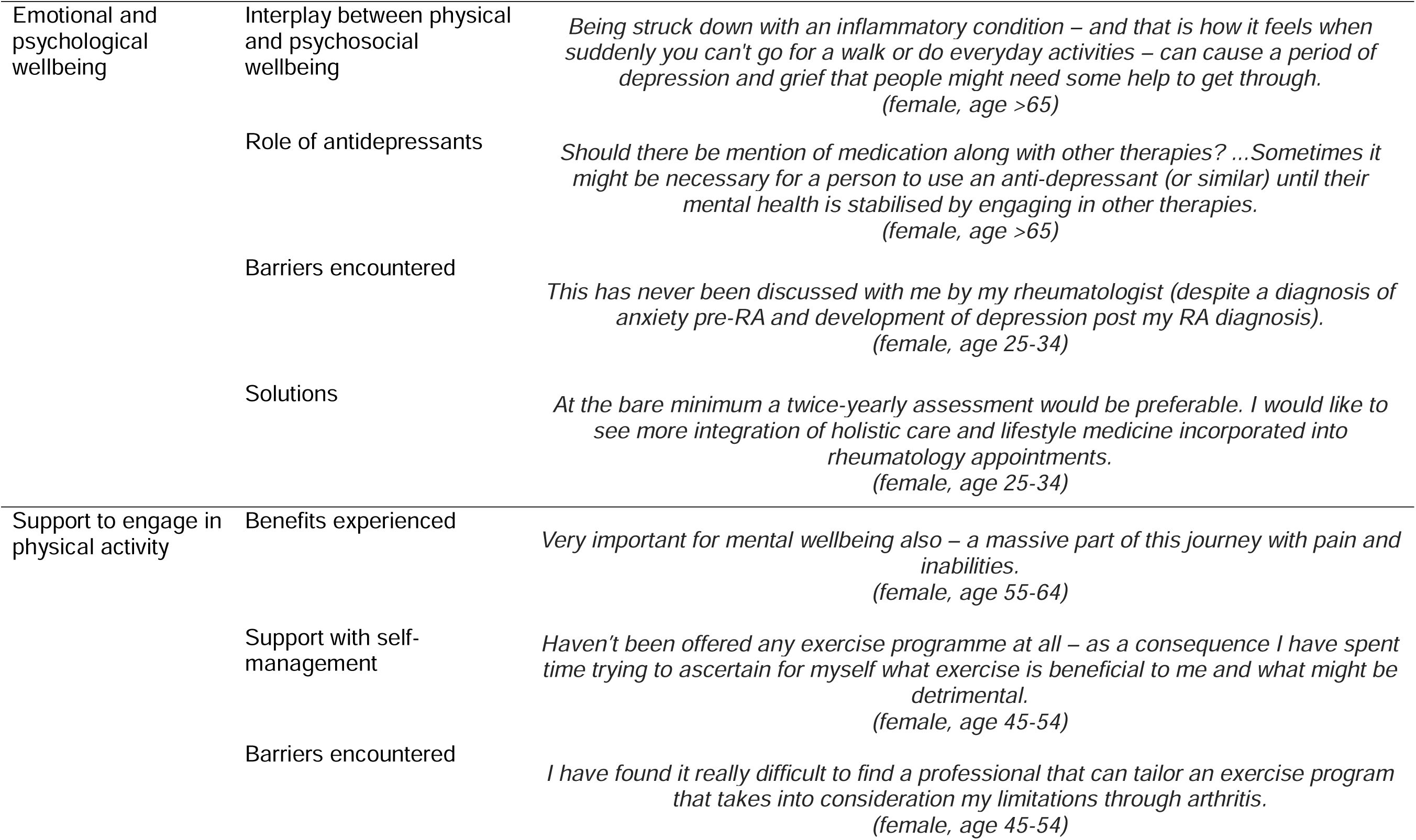
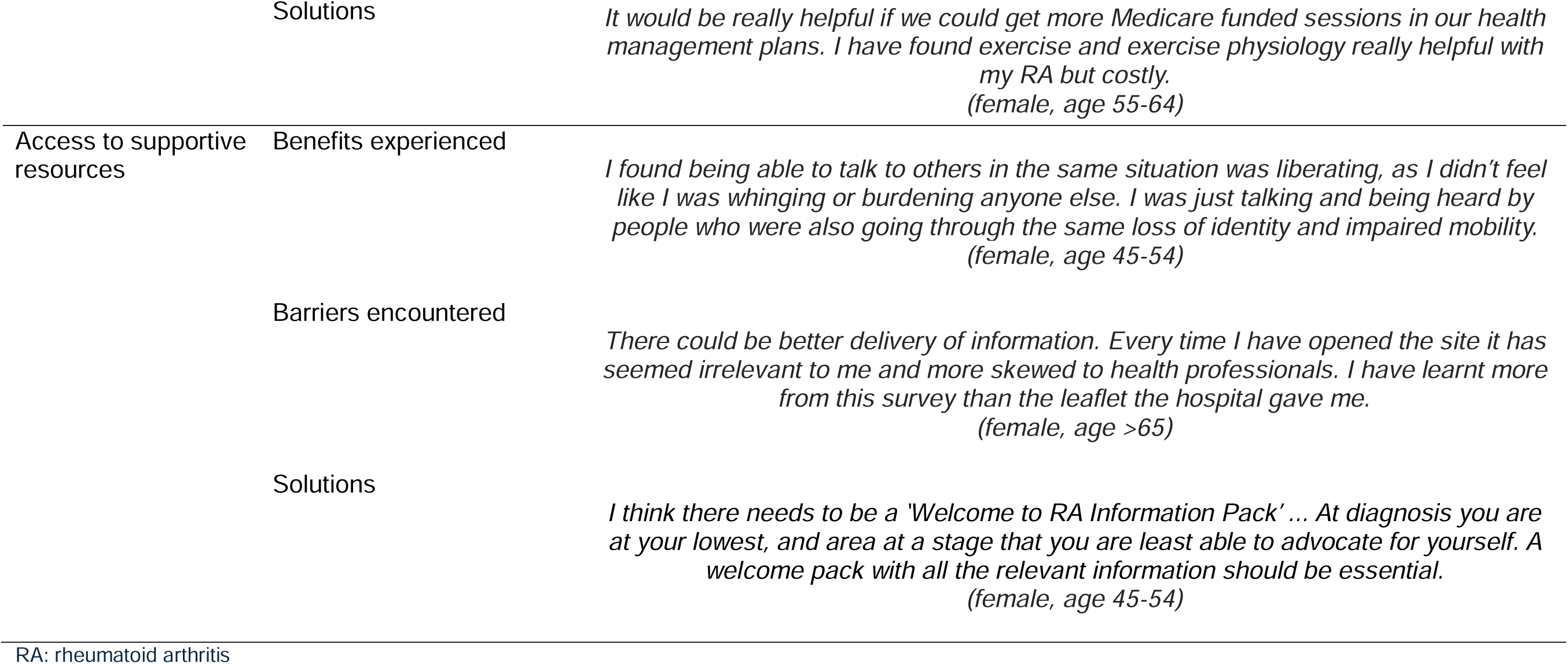
Thematic analysis for supportive care quality statements.

The complexity of pain management was also discussed, and issues with both pharmacological and holistic approaches were highlighted. Consumers believed analgesia was necessary to improve function and quality of life but also disliked being reliant on medications, especially due to the stigma surrounding opioid use and the side effects they experienced. Some consumers resented the recent shift towards holistic and mindfulness-based pain management, viewing it as a barrier to accessing necessary medications. However, others found these approaches useful as complementary or alternative therapies.

Consumers identified several barriers to effective pain management including high cost, insufficient services, and limited knowledge about RA amongst care providers. Difficulties in accessing pain medication, particularly due to government restrictions on opioid prescriptions and poor healthcare professional understanding, further impacted pain management. Consumers also struggled with self-management and wanted more professional guidance, including the development of self-help tools and ‘action plans’ for patients to refer to.

##### EMOTIONAL AND PSYCHOLOGICAL WELLBEING

The complex interplay between physical health and mental or emotional wellbeing was a key theme raised by consumers. Pain and impaired function contributed to depression, isolation, stress on relationships, and difficulties with employment. Emotional and psychological support was seen as beneficial for improving pain levels, mental health, and social wellbeing. However, some consumers raised concern that an overemphasis on mental wellness programs made them feel dismissed and created barriers to accessing pain relief. Conflicting views on antidepressants were also discussed; some believed they were useful adjuncts to alternative therapies, whereas others felt they were ineffective or used to avoid exploring other management options.

Barriers to support at a systems level included a lack of appropriate services, the complexity of care coordination, and high costs of multidisciplinary care. At the healthcare level, consumers expressed frustration with assessments that focused primarily on physical health and did not address mental and emotional wellbeing, even for those with pre-existing mental health issues. Mental health was difficult for consumers to raise with their healthcare providers, as they felt these concerns were not taken seriously. Consumers suggested that increased funding and frequent assessments, at all stages in their disease course, were required to ensure access to support when needed.

##### SUPPORT TO ENGAGE IN PHYSICAL ACTIVITY

Consumers reported several benefits of physical activity, including improved pain control, function, and independence. Furthermore, the social interaction offered by physical activity was felt to enhance mental wellbeing and reduce isolation. Some consumers felt unsupported in engaging with physical activity, and personalised advice was needed to accommodate physical limitations, avoid flares, and improve understanding of the benefits of physical activity.

Challenges faced by patients involved psychological barriers, such as low confidence or fear of injury, and physical limitations from RA, comorbidities, or pain. Other barriers included high cost, limited available services, lack of awareness of resources, and a shortage of specialised advice. Consumers suggested increasing funding, training opportunities, and government subsidies to address these issues. Consumers also suggested services and resources they felt would be beneficial; these included specialised fitness facilities, programmes, and self-help resources such as exercise videos tailored to RA patients.

##### ACCESS TO SUPPORTIVE RESOURCES

Supportive resources empowered patients by increasing their understanding about RA and the available services. Peer support services also helped to alleviate isolation and improve knowledge. However, many consumers felt that these resources should not be viewed as all-encompassing and wanted additional explanations from the healthcare team. Some also felt that supportive resources were not able to provide practical management solutions.

Barriers to accessing supportive resources included lack of referral and awareness, inadequate quality of available resources, technological and proximity issues, and scheduling conflicts. Consumers suggested ways to improve the format and content of resources, methods to supplement resources and increase their utility, and ideas for placement of resources to improve access.

#### THEMATIC ANALYSIS FOR QUALITY STATEMENTS RELATED TO MEDICAL MANAGEMENT (Table 4)

##### ACCESS TO SPECIALIST CARE

Consumers frequently reported barriers to accessing specialist RA care. Delays in diagnosis occurred due to symptoms being dismissed, atypical presentations, inappropriate investigations, or normal inflammatory markers and seronegative results causing confusion. Primary care limitations further delayed referrals to specialists, and insufficient specialist services, particularly in rural areas, led to lengthy waiting periods and the necessity of long-distance travel. Timely treatment was also hindered by difficulty accessing rheumatologists, and general practitioners (GPs) often lacked the confidence to initiate RA management without specialist input.

**Table 4:**
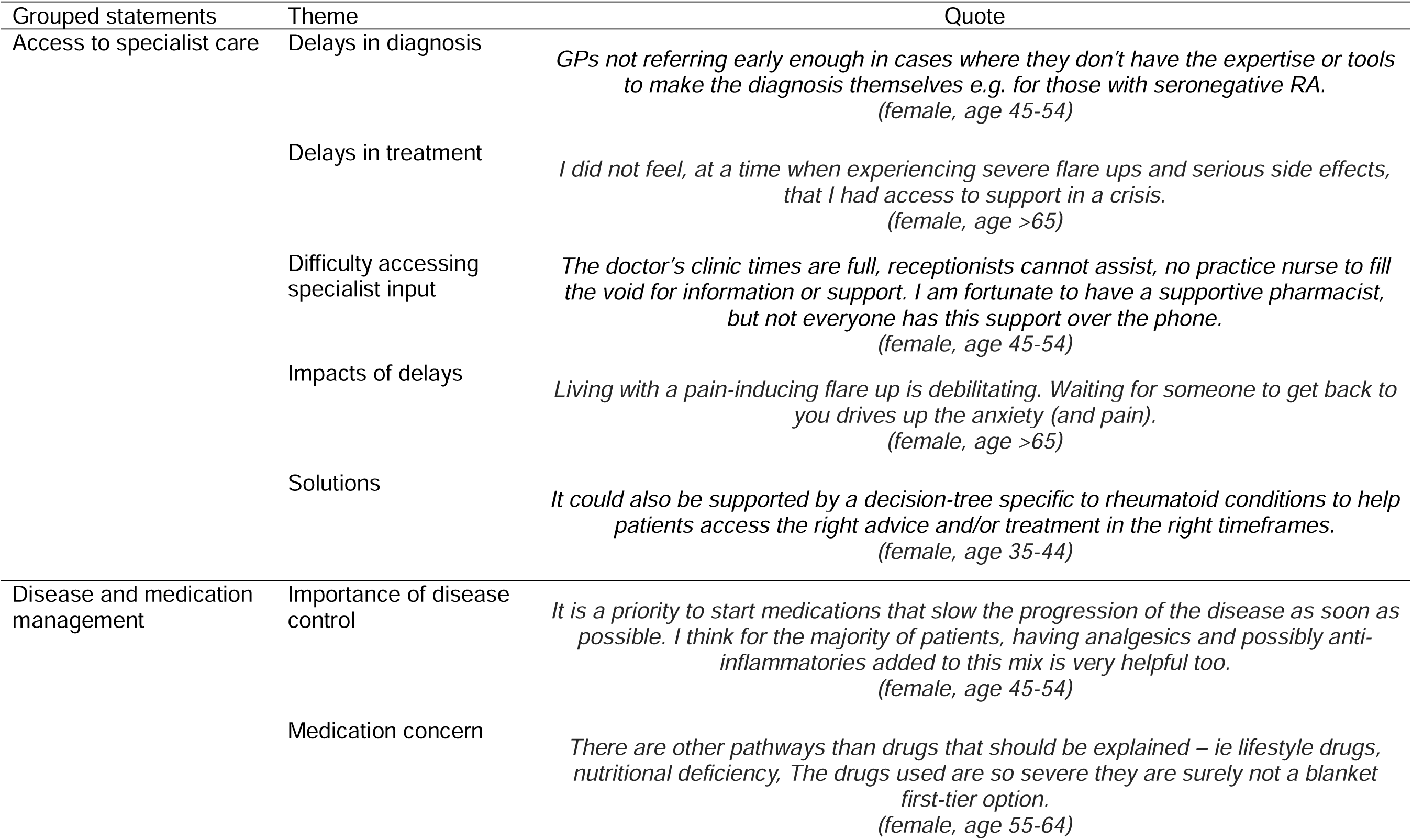
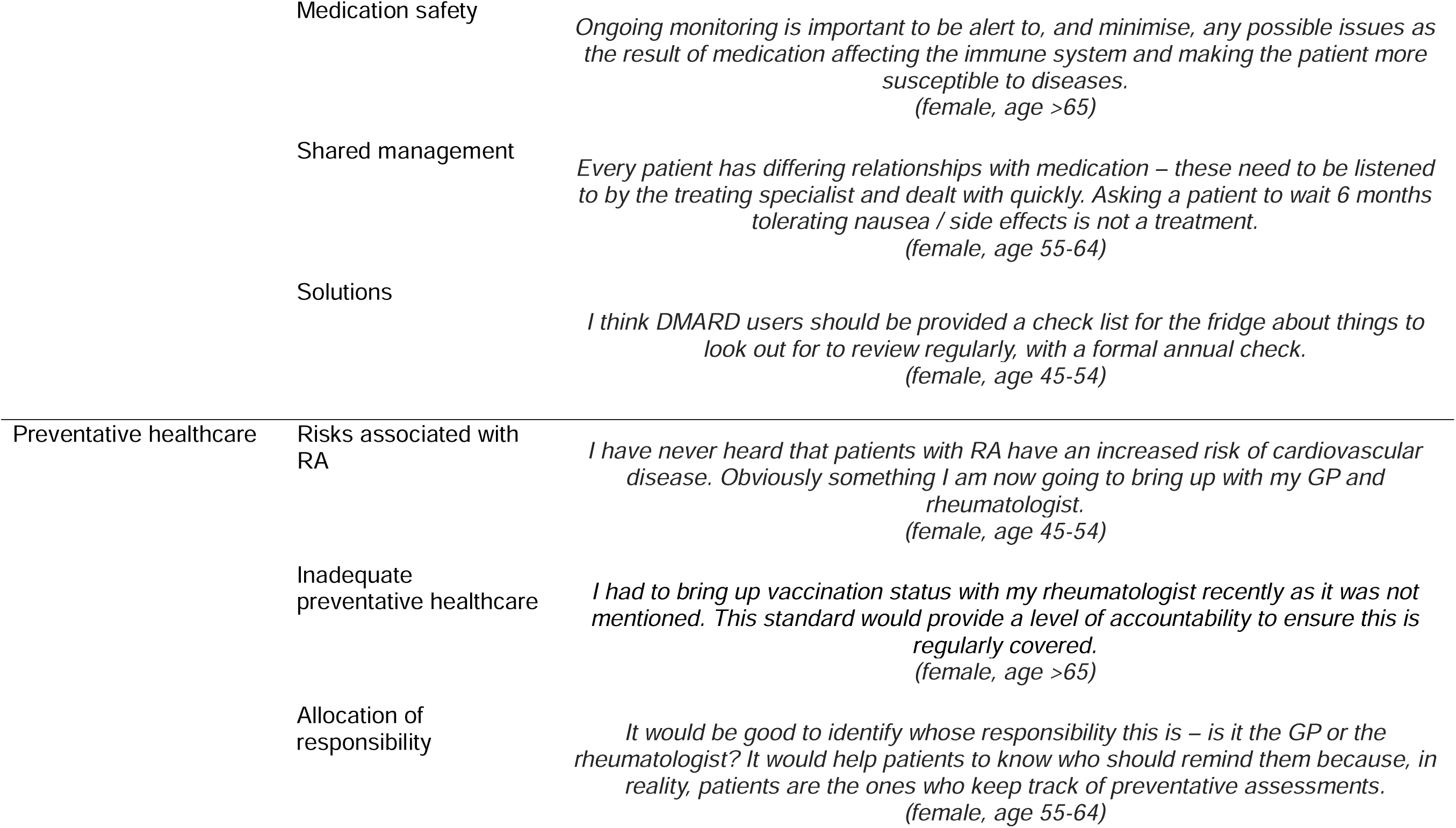
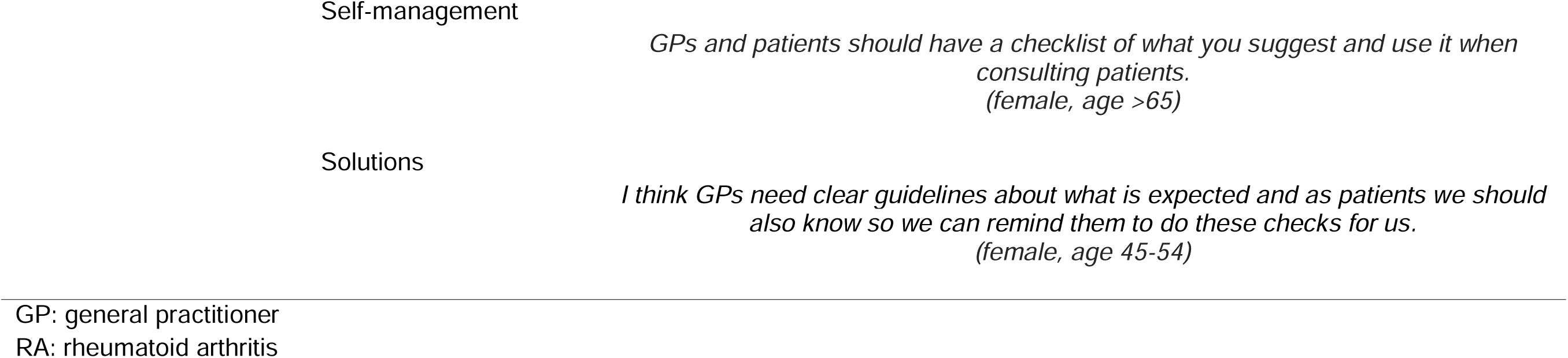
Thematic analysis for grouped quality statements.

Patients struggled to access interim specialist advice between scheduled visits, noting that GPs and other care providers frequently lacked sufficient RA-specific knowledge to adequately support them. Consequently, patients experienced anxiety, unmanaged pain, functional impairment, medication-related issues, and sometimes resorted to self-management without professional guidance. Consumers suggested improvements such as GP education regarding RA symptom recognition and referral processes, establishment of rheumatology telephone support lines, involvement of other appropriately trained healthcare providers, and development of self-management resources and ‘action plans.’

##### DISEASE AND MEDICATION MANAGEMENT

The importance of disease control was frequently emphasized by consumers, with most believing that early and effective treatment is important to prevent damage and improve quality of life. Poor disease control was seen to negatively impact pain, function, social participation, and emotional wellbeing. Barriers to early treatment included diagnostic uncertainty, inability of GPs to initiate treatment, long wait times for rheumatology input, the need for safety checks, and patient concerns about medications. Consumers also described difficulty in establishing an effective treatment plan, needing to cycle through multiple medications before achieving disease control.

Consumers had varying views on DMARD therapy, with some advocating for immediate access to biologics whilst others preferred more time to make informed decisions and consider alternative or nonpharmacological measures first. Several medication concerns were discussed and cited as reasons for patients to avoid starting treatment. Medication counselling was felt important to alleviate concerns, improve understanding, and foster engagement with proposed treatment plans. Consumers valued medication safety, including necessary safety checks and monitoring for adverse effects.

Specialists were expected to provide up-to-date and evidence-based management plans, but also to individualise the approach based on patient comorbidities, personal goals, and preferences. Emphasis was given to the importance of developing shared management plans with patients which incorporate biochemical, functional, and psychosocial goals.

Consumers also valued self-management and provided several practical suggestions to support this, including educational resources and checklists for the fridge.

##### PREVENTATIVE HEALTHCARE

Many consumers reported poor understanding about the increased health risks associated with RA and were unaware of preventative measures they could take. Some discussed the interaction of RA with other risk factors, such as physical limitations contributing to falls and fracture risk or medications contributing to infections. Confusing advice from healthcare professionals and lack of risk assessment were felt to hinder preventative healthcare. Consumers suggested that regular and individualised risk assessments, supported by RA-specific guidelines, could improve preventative care.

Confusion about the allocation of responsibility for preventative health was evident. Although most consumers believed their GP should be responsible for assessing and managing preventative health, many were concerned that they lacked understanding of the broader health issues related to RA. Some consumers relied on their rheumatologist for preventative health measures, but most saw their role as supportive and expected them to provide advice to both GPs and patients. Consumers described feeling unsupported and overwhelmed with preventative health and, once again, guidelines or checklists were suggested as helpful tools to empower patients in this area.

## DISCUSSION

This study provides important insight into the challenges in RA management faced by consumers, as well as potential approaches to improving the quality of care received. Our study highlights the broad impacts of RA on physical, mental, emotional, and social wellbeing. Consumers stressed the importance of patient involvement in care and felt that pain management, function, and psychosocial wellbeing were often not prioritised. A range of barriers were identified across the availability, accessibility, and suitability of care in RA.

### Impacts of RA

Consumers identified impacts of RA on function, mobility, social participation, independence, employment, and mental wellbeing. Pain and disease activity were identified as major drivers of these impacts. Previous studies have also demonstrated that physical wellbeing and disease burden influence psychological, social, and occupational aspects of patients’ lives^12,13^. Consumers felt controlling disease activity and symptoms were important for quality of life, but they also cited concern about the negative impacts of treatment. Fear of medication, maintaining a sense of control over health, disappointment with treatment, and feeling overwhelmed with treatment decisions have previously been identified as motivations for resisting DMARD therapy in RA^14^. Consumers in our study reported debilitating side effects from DMARD therapy, and fear of complications caused some to delay starting treatment. Stigma or shame around the use of pain medication was also experienced.

Consumers stressed the value of medication counselling, education for patients and healthcare providers, and developing shared goals to support patients and mitigate these issues.

### Patient involvement

Patient involvement in care, through shared decision-making and self-management strategies, was another key theme in our study. Consumers wanted to be actively involved in their care but reported feeling unsupported and isolated. Feelings of being dismissed, or not having their concerns and management priorities addressed, were frequently reported. Consumers felt that current models of care did not capture the many facets of RA, focusing more on physical health without addressing social and emotional wellbeing. Active patient involvement in care improves acceptability of and adherence to management plans in RA, ultimately improving patient outcomes and satisfaction^6,8,12,13^. Current international practice guidelines highlight the importance of active participation and self-management for patients with RA^3,7,15^. Several practical suggestions to empower self-management were offered by consumers in our study; this included checklists and guidelines for preventative care and medication monitoring, ‘action plans’ for pain and flare management, and self-guided exercise programmes.

### Barriers identified and potential solutions

#### Availability

Consumers identified issues with availability of care, including a lack of specialised health practitioners. Prolonged wait times, delays in diagnosis and treatment, and difficulties in accessing appropriate advice were reported. This resulted in worsening symptomatology and impacts on mental health. These issues are reflective of the current workforce shortage in Australia amongst rheumatologists, as well as rheumatology-specific nursing and allied health staff ^16,17^. Multidisciplinary care is known to improve patient outcomes and is considered an essential part of RA management, and nurse-led RA interventions have been shown to improve the quality and efficacy of care delivery^18–21^. In addition to training more rheumatologists, consumers in our study suggested that upskilling other health professionals to provide specialised support alongside rheumatologists would be valuable.

#### Accessibility

Participants reported accessibility of healthcare as another barrier. Prohibitive cost and insufficient funding were prominent themes, particularly for multidisciplinary care and accessing allied health services. A recent study evaluating the nursing and allied health workforce in Australian public rheumatology departments also identified cost to be the most common barrier^17^. At present, Medicare funding is available in Australia to subsidise up to five allied health visits per calendar year, and out-of-pocket healthcare costs for people living with RA are substantial^22,23^. Consumers in our study frequently referenced that current funding is insufficient and suggested that specific additional subsidies for people living with RA would be of benefit.

Other issues with accessing care included a lack of referral to, or awareness of, recommended services and resources. This was experienced for support groups, multidisciplinary care, and preventative health. Issues in referral pathways have previously been identified at the GP level, but also in nursing and allied health settings^17,18^. Consumers suggested that clearer referral pathways, allocation of responsibility for care, and high-quality resources for patients and GPs would help to overcome some of these barriers.

Rural consumers experienced more difficulty in accessing specialist and supportive care compared to those in urban areas. This is reflective of the relative concentration of healthcare services in metropolitan Australia^16,24^. Telehealth consultations, self-help resources, and upskilling of GPs were amongst the suggestions offered by consumers to improve care accessibility.

#### Misalignment of services

Consumers felt there was a paucity of providers with RA-specific knowledge, resulting in conflicting or ill-informed advice. Personalised care and specialised advice were wanted, the benefits of which have previously been recognised^19,20^. Upskilling allied health professionals and developing RA-specific services or resources, such as fitness facilities and self-help tools, were suggested as ways to improve the quality of care received.

Improvement at the primary care level was also desired, and consumers felt that GPs did not have enough knowledge about RA to provide the support patients need. Consistent with previous studies, this was identified as an issue across diagnosis, initial management, flare management, pain management, and preventative healthcare^18^. Education, clinical resources, and development of guidelines were suggested as ways to support GPs, and the importance of communication between specialists and GPs was also stressed.

#### Limitations and strengths

There are several limitations to our study, including the cross-sectional nature and the potential for selection bias given recruitment of participants was limited to people already registered with arthritis organisations. Although a survey-based approach may limit the depth of data collected from individuals, it allowed for large-scale sampling of RA consumers nationally which would not have been possible through other qualitative methods, such as interviews or focus groups. Strengths of our study included the national recruitment and large sample size. Furthermore, consumer involvement was sought throughout the course of our study, with their input contributing to the development of the standard as well as in thematic analysis and co-authorship. This strengthened our study design and improved the relevance of our findings.

### Conclusions

Our study explored current consumer experiences of RA management in Australia and provides important insight into consumer care priorities, which will inform the development of RA quality care indicators. We identified key impacts of RA experienced by patients, including the interplay between physical and psychosocial wellbeing. Consumers encountered significant barriers to care including issues with availability, accessibility, and suitability of services. Several practical solutions were offered, which can be used to inform practice and empower patients in the management of RA.

Our findings are reflected in the guiding principles underpinning the RA Clinical Care Standard and demonstrate that consumers value holistic and team-based care, effective communication, shared decision-making, and self-management.

## Data Availability

All data produced in the present study are available upon reasonable request to the authors.

## ACKNOWLEDGEMENTS

The authors thank the consumers participating in the survey. We also thank Dr Elizabeth Hoon for her guidance and support in the approach to qualitative analysis, and the ARA RA Clinical Care Standard Working Group involved in the development of the Standard.

## AUTHOR CONTRIBUTIONS

All authors contributed to study conception and design, as well as analysis and interpretation of data. Data acquisition was performed by MS. All authors have reviewed and critically revised the manuscript and approved the final version for publication.

## Statement of ethics and consent

Ethical approval for Working Group participation and survey participation was obtained from Central Adelaide Local Health Network Human Research Ethics Committee (No. 17784). This study involved an anonymous online survey, and no identifiable data was collected. The purpose of the study, as well as information about data use and the voluntary nature of participation, was outlined on the introductory page. Informed consent was implied through completion of the survey.

